# Inequalities in physical fitness in children with hearing loss: a systematic review and meta-analysis with implications for inclusive physical education and school curriculum

**DOI:** 10.64898/2026.06.08.26355131

**Authors:** María Victoria Diaz-Franco, Alexis Caniuqueo-Vargas, Olusiji Adebola Lasekan, Carlos Alberto Castillo-Sarmiento, Beatriz Rodríguez-Martín

## Abstract

**Background:** Childhood and adolescent hearing loss affects not only communication and cognitive development but also motor skills and school participation. Consequently, it generates inequalities in learning and educational inclusion. Nevertheless, no systematic review has yet analyzed these differences from an inclusive education perspective.

**Methods:** A systematic review with meta-analysis was conducted following PRISMA guidelines and registered in PROSPERO. Observational studies comparing physical fitness between children and adolescents with hearing loss and their hearing peers were included. Methodological quality was assessed using the Newcastle–Ottawa Scale, and standardized effect sizes were calculated with a random-effects model.

**Results:** Five studies (*n* = 404) were analyzed. Findings revealed significant differences in strength, agility, speed, and balance. Moreover, the meta-analysis showed a large standardized effect favoring hearing children (ES = −2.35; 95% CI: −3.34 to −1.37).

**Conclusions:** Children and adolescents with hearing loss present significantly lower physical fitness, which may affect the planning of physical education activities if their capacities are misinterpreted. Implementing inclusive and adapted strategies within the school curriculum is essential to ensure equal opportunities, improve physical fitness, and promote educational equity.

## 1. Introduction

Hearing loss (HL) is defined as a reduction in auditory capacity in both ears exceeding 20 decibels (dB). According to the World Health Organization (WHO), more than 5% of the global population requires rehabilitation for HL, including 34 million children (World Health Organization, 2026). The main causes include otitis media, otosclerosis, meningitis, ototoxic medications, head trauma, genetic disorders, acoustic trauma, and noise exposure (Morros-González et al., 2022). In addition, lifestyle factors such as diabetes (ALDajani et al., 2015), obesity (Shafiepour et al., 2022), and metabolic syndrome (Rim et al., 2021) have also been associated with HL.

These conditions reduce auditory input and generate weak signals that affect information processing. As a result, sensorimotor development is limited (Blank et al., 2012; De Kegel et al., 2011). For instance, children with HL often present communication difficulties, learning challenges, and social interaction problems (Roland et al., 2016). They also experience psychomotor delays, motor coordination and balance disorders (Blank et al., 2012; De Kegel et al., 2011). Moreover, studies have reported delays in motor skill acquisition (Dummer et al., 1996; Slavnić et al., 2017; Weiss and Phillips, 2006) and coordination difficulties (Melo et al., 2012; Zwierzchowska et al., 2020).

Physical fitness (PF) is not only a predictor of health (Martin et al., 2014; Recchia et al., 2023) but also a preventive and therapeutic mechanism for various diseases. However, PF is reduced in children with HL, producing negative effects on overall development (Healy et al., 2020; Novikov et al., 2019; Ortega et al., 2025; Shavel et al., 2021; Zwierzchowska et al., 2020). PF variables such as body composition, muscular strength, endurance, cardiorespiratory fitness, and flexibility are associated with better health outcomes and academic performance (Abrantes et al., 2022; Martin et al., 2018). Therefore, PF development during childhood and adolescence is not only a predictor of adult health; it is also a key factor for school participation and educational inclusion (Recchia et al., 2023; Rojas-Rueda et al., 2021).

Recent literature highlights that inequalities in PF may limit full participation in the school curriculum and create barriers to equity. Escriva-Boulley et al. (Escriva-Boulley et al., 2021) emphasize that inclusive teaching practices in physical education require institutional and methodological support. Moreover, Bailey and Sweeney (Bailey and Sweeney, 2022) identified principles and strategies of inclusive physical activity that guide teachers in adapting content and methodologies. In particular, Escriva-Boulley et al. (Escriva-Boulley et al., 2021) argue that inclusive physical education (PE) is essential to promote equity and participation in schools.

Therefore, the aim of this study was to conduct a systematic review and meta-analysis to determine differences in PF variables between children and adolescents with HL and their hearing peers. The purpose was to provide evidence to inform teaching practice, inclusive curricula in physical education, and adapted pedagogical strategies, thereby promoting educational equity.

## 2. Methods

A systematic review and meta-analysis was conducted following the principles of the PRISMA statement (Hutton et al., 2016) and in accordance with the Cochrane Collaboration Handbook (Higgins et al., 2024).

The protocol of this review was registered in PROSPERO (CRD42024593329). Peer-reviewed articles were identified through searches in PubMed, Scopus, and Web of Science databases, from inception to December 2025 (see Supplementary Material 1 for the search strategy). Ethical approval and informed consent were not required; however, the authors declare compliance with international research codes, including the Declaration of Helsinki. To verify the quality of the search strategy, two reviewers (AC, VD) independently identified peer-reviewed articles, with disagreements resolved by a third reviewer (BR).

### 2.1. Inclusion and exclusion criteria

Studies were included if they met the following criteria: (1) observational analytical or case-control studies; (2) participants were school-aged children and adolescents; (3) publications in English, Spanish, or Portuguese; (4) studies including a diagnosis of HL and performance in PF tests; and (5) studies reporting mean and standard deviation values of performance variables. Exclusion criteria were systematic reviews, meta-analyses, studies without a hearing control group, qualitative studies, and grey literature (e.g., book chapters, opinion articles), as they did not provide sufficient data for analysis. The PRISMA flow diagram (Figure 1) details the study selection process.

**Figure 1:**
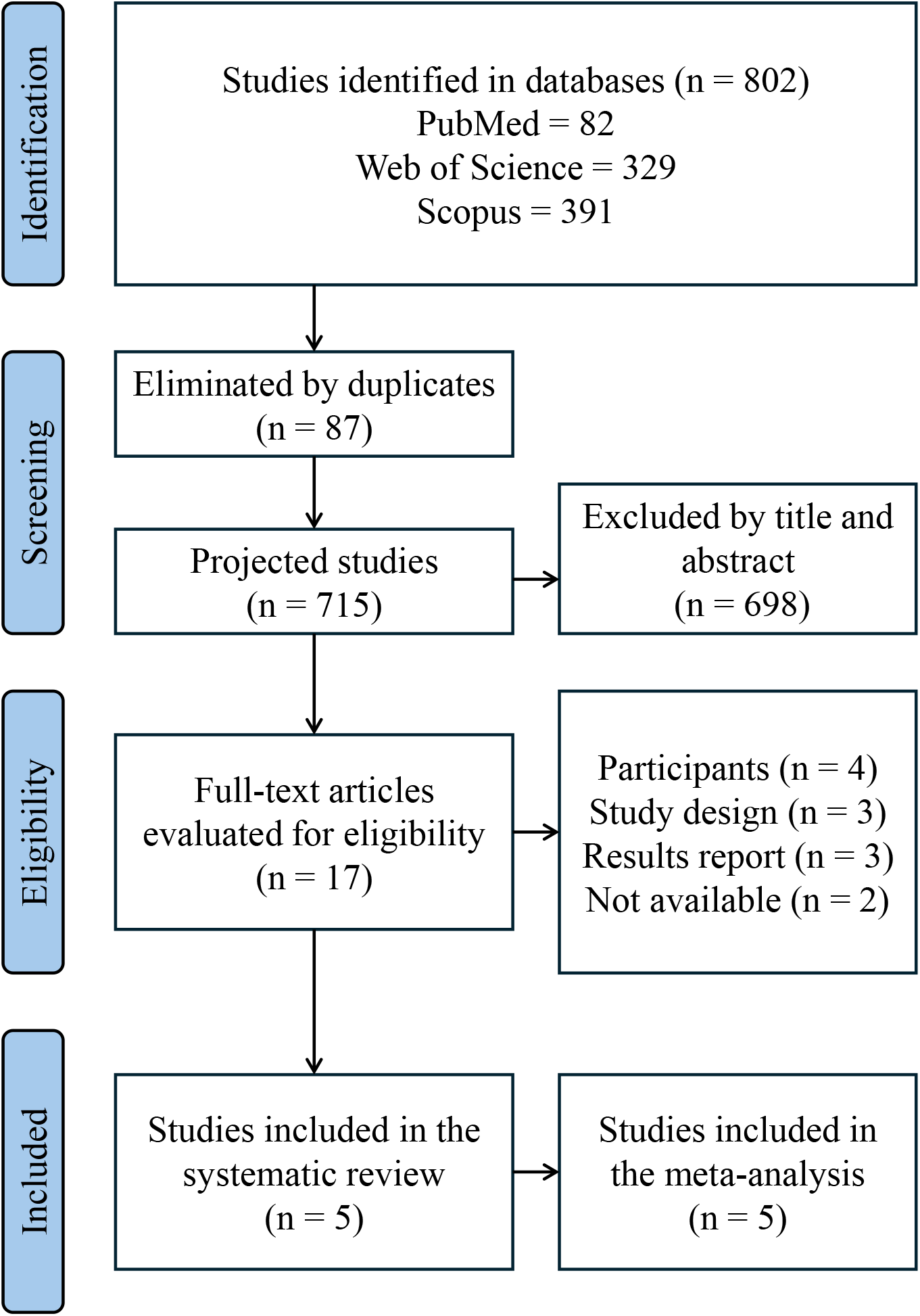
PRISMA flow diagram illustrating the identification, screening, eligibility assessment, and inclusion of studies in the systematic review and meta-analysis.

### 2.2. Data extraction

Two reviewers (VD, AC) independently extracted data into a predesigned Excel sheet (Supplementary Material 2). Only exact information was extracted; if data were not adequately reported, they were documented as “not indicated”. Disagreements were resolved by the third reviewer (BR). Extracted data included: first author, year and country, study design, number of participants, sex, age, PF criteria, HL criteria, groups with and without HL, instruments used to assess PF and HL, mean and standard deviation of performance results, and study conclusions.

### 2.3. Risk of bias and methodological quality

Risk of bias and methodological quality were assessed using the Newcastle– Ottawa Scale (NOS). The NOS evaluates three domains: selection, comparability, and outcome/exposure. Scores of 0–4, 5–6, 7–8, and 9–10 correspond to low, moderate, high, and very high quality, respectively. Only studies of high and very high quality were included. Results of the quality assessment are presented in Table 1.

**Table 1:** Methodological quality assessment of the included studies using the Newcastle– Ottawa Scale (NOS).

| Study | C1 | C2 | C3 | C4 | Comp. | E1 | E2 | E3 | Score |
| --- | --- | --- | --- | --- | --- | --- | --- | --- | --- |
| Azadian (2020) | * | * | * | * | * | * | * | * | 8 |
| Banerjee (2022) | * |  | * | * | ** | * | * | * | 8 |
| Majlesi (2014) | * | * | * | * | * | * | * | * | 8 |
| Melo (2012) | * | * | * | * | * | * | * | * | 8 |
| Shavel (2021) | * |  | * | * | * | * | * | * | 7 |

C1: Is the case definition adequate?; C2: Representativeness of the cases; C3: Selection of controls; C4: Definition of controls; Comp.: Comparability of cases and controls on the basis of design or analysis; E1: Ascertainment of exposure; E2: Same method of ascertainment for cases and controls; E3: Non-response rate.

### 2.4. Statistical analysis

Standardized mean effect sizes (ES) were calculated between groups with and without HL using Hedges’ *g* (Hedges and Olkin, 2014). Effect sizes were interpreted as small (*<* 0.20), medium (0.20–0.50), moderate (0.50–0.80), and large (*>* 0.80) according to Cohen (2013). A random-effects model was applied.

Heterogeneity was assessed using *Q* and *I*^2^ statistics. For *Q, p <* 0.1 was considered statistically significant. For *I*^2^, cut-off points were *<* 25% (low), 25–50% (moderate), *>* 50–75% (high), and *>* 75% (severe) heterogeneity (Deeks et al., 2019). Chi^2^ tests were used to determine whether observed differences were consistent with proportions.

Publication bias was evaluated using funnel plot interpretation (Zwetsloot et al., 2017). Quantitative analyses were performed using Cochrane Review Manager software (RevMan version 5.1, Cochrane IMS, Denmark).

## 3. Results

The database search identified 802 potential articles. Of these, 87 were removed as duplicates. Another 704 were excluded for not meeting the inclusion criteria. Twelve full-text articles were excluded for lacking a hearing control group. Finally, five high- and very high-quality studies were included in the systematic review and meta-analysis, with a total of 404 participants (see Figure 1).

### 3.1. Strength variables

Two studies evaluated strength as an indicator of PF (Azadian et al., 2020; Shavel et al., 2021). Azadian and colleagues (Azadian et al., 2020), in a sample of 30 children (15 with HL and 15 hearing), assessed hip, knee, and ankle torque using force plates and motion capture. Children with HL showed lower joint torques in ankle evertors, knee flexors, and hip abductors compared to hearing peers (*p <* 0.05). In the dominant limb, maximum torques in ankle dorsiflexion, knee adduction, and hip adduction were significantly higher in hearing children (*p <* 0.001). Shavel et al. (Shavel et al., 2021), in a sample of 94 children (72 with HL and 22 hearing), used the PWC150 test and found reduced performance in children with HL across both sexes (*p <* 0.05).

### 3.2. Agility and speed

Two studies assessed agility and speed (Banerjee et al., 2022; Majlesi et al., 2014). Banerjee and co-authors (Banerjee et al., 2022), with 252 children (42 with HL and 210 hearing), applied the 4 × 10 m shuttle run test. Results showed significant differences (*p <* 0.05), with agility increasing as HL severity decreased. Majlesi and colleagues (Majlesi et al., 2014), in a sample of 20 children (10 with HL and 10 hearing), reported higher walking speed in hearing children.

### 3.3. Balance

Two studies evaluated balance (Majlesi et al., 2014; Melo et al., 2012). Majlesi and colleagues (Majlesi et al., 2014), in the same sample of 20 children, found greater sway in children with HL compared to hearing peers (*p <* 0.05). They concluded that exercise programs may improve somatosensory capacity and balance. Melo and colleagues (Melo et al., 2012), with 88 children (44 with HL and 44 hearing), assessed balance and gait using the Tinetti Balance and Mobility Scale and the Timed Up and Go test. Hearing children obtained higher scores in both domains, with particularly marked differences in gait performance (*p <* 0.001).

### 3.4. Meta-analysis

The meta-analysis revealed a large standardized effect size (ES = *−*2.35 [95% CI: *−*3.34 to *−* 1.37], *Z* = 4.68, *p* = 0.00001), indicating reduced PF in children and adolescents with HL compared to hearing peers. However, the small number of studies limited subgroup analyses, as authors did not stratify by age, degree of HL, or other design characteristics. Consequently, heterogeneity was severe (*I*^2^ = 92%, *p* = 0.00001), suggesting substantial variability across studies. Owing to the limited number of included studies, subgroup analyses according to age, severity of hearing loss, or other study characteristics could not be performed. Detailed meta-analytic results are presented in Table 2, and the corresponding forest plot is shown in Figure 2.

**Table 2:** Summary of studies included in the meta-analysis and standardized effect sizes comparing participants with hearing loss and hearing controls.

| Study | Hearing loss |  |  | Control |  |  | Weight (%) | SMD (95% CI) |
| --- | --- | --- | --- | --- | --- | --- | --- | --- |
|  | Mean | SD | n | Mean | SD | n |  |  |
| Azadian (2020) | 0.53 | 0.21 | 15 | 0.65 | 0.12 | 15 | 16.9 | -0.68 [-1.42, 0.06] |
| Banerjee (2022) | 14.01 | 0.93 | 42 | 11.39 | 0.87 | 42 | 17.4 | -2.88 [-3.50, -2.26] |
| Majlesi (2014) | 4.90 | 0.46 | 10 | 3.23 | 0.46 | 10 | 13.1 | -3.48 [-4.97, -1.99] |
| Melo (2012a) | 13.60 | 1.00 | 44 | 14.80 | 0.90 | 44 | 18.0 | -1.25 [-1.71, -0.79] |
| Melo (2012b) | 9.90 | 1.30 | 44 | 11.80 | 0.50 | 44 | 17.8 | -1.91 [-2.42, -1.40] |
| Shavel (2021) | 301.45 | 2.77 | 72 | 312.72 | 2.04 | 22 | 16.7 | -4.27 [-5.05, -3.48] |
| Overall |  |  | 227 |  |  | 177 | 100.0 | -2.35 [-3.34, -1.37] |

Random-effects model. Heterogeneity: $\tau^2 = 1.35$ ; $\chi^2 = 66.18$ , $df = 5$ ( $p < 0.00001$ ); $I^2 = 92\%$ . Test for overall effect: $Z = 4.58$ ( $p < 0.00001$ ). Negative standardized mean differences indicate lower physical fitness performance among participants with hearing loss.
Melo (2012a) corresponds to the balance subscale and Melo (2012b) to the gait subscale of the Tinetti Balance and Mobility Scale.

**Figure 2:**
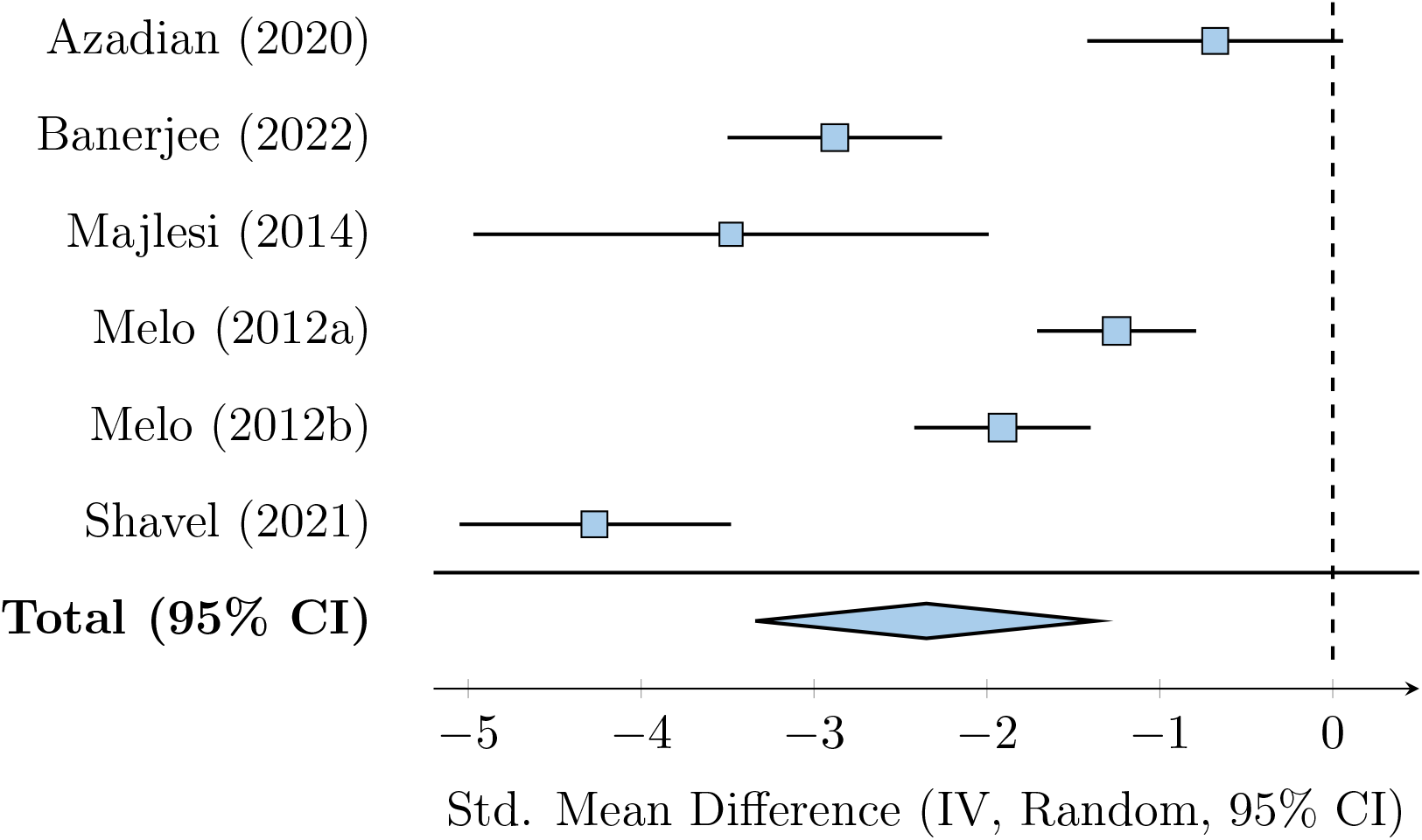
Forest plot showing the standardized effect sizes comparing physical fitness outcomes between children and adolescents with hearing loss and hearing controls.

## 4. Discussion

This meta-analysis confirms that children and adolescents with HL perform worse in PF. Strength, agility, speed, and balance were consistently lower than in hearing peers. These findings highlight a clear gap that teachers must consider when planning PE. In addition, it has been reported that individuals with HL present poor physical and psychological performance, along with reduced capacities for social and cognitive development (Martinez-Amezcua et al., 2021), emotional and neuromotor development (Dammeyer, 2010; Dammeyer et al., 2018; Hogan et al., 2014), decreased muscle tone (Melo et al., 2012, 2021; Yamazaki et al., 2023), reduced ability to maintain balance, and difficulties in performing complex coordinated movements (Neszmélyi and Horváth, 2019; Slavnić et al., 2017; Stepanchenko et al., 2020).

Regarding strength, our results revealed lower performance in hip, knee, and ankle torque, consistent with findings reported by Azadian and colleagues (Azadian et al., 2020) and Shavel and colleagues (Shavel et al., 2021). Differences were also observed in running speed, walking speed, and balance tests (Banerjee et al., 2022; Majlesi et al., 2014; Melo et al., 2012). These inequalities in physical performance appear to be related to lower levels of physical activity among children with HL (Abi Nader et al., 2018; An, 2019; da Silva et al., 2019; Godoy-Cumillaf et al., 2022). Consequently, reduced participation in physical activity may partially explain the lower levels of physical fitness observed in children and adolescents with HL.

In addition, reduced social participation, weaker psychosocial skills, and lower self-confidence in children with hearing loss negatively affect quality of life and school participation (Aggarwal et al., 2024; Roland et al., 2016).

These inequalities go beyond biomedical explanations and must be understood in the context of PE. Physical activity is a central element for equitable participation in schools. Recent European research shows that inclusion in PE is still limited and often depends on institutional support and teacher training (Marron et al., 2023). Evidence also indicates that the presence of students with special needs is not enough; teachers must adapt both content and methodology. In Spain, for example, structural barriers and the demand for specialized training remain significant (Saiz-González et al., 2025). Other studies stress that effective inclusion requires institutional backing and teacher preparation (Escriva-Boulley et al., 2021). Principles and strategies for inclusive physical activity have been proposed to guide curricular adaptation (Bailey and Sweeney, 2022). Finally, the literature highlights the complexity of inclusion, which involves overcoming cultural and structural barriers to ensure equal participation (Escriva-Boulley et al., 2021).

Other studies confirm that inclusion in physical education remains a challenge. Despite progress, barriers related to resources, teacher training, and attitudes toward disability persist (Morley et al., 2021). These findings rein-force the need for programs that explicitly incorporate inclusion and accessibility into the curriculum. This approach aligns with UNESCO recommendations, which highlight PE as a low-cost, high-impact strategy to promote equity and public health (Miao et al., 2021).

Furthermore, inclusion should not be limited to individual adjustments. It must be embedded in an inclusive curriculum that integrates cooperative learning, visual supports, differentiated instruction, and adapted assessment (Karamani et al., 2024). A recent systematic review confirms that strategies such as co-teaching and peer tutoring are effective in improving inclusion and socialization in physical education (Ben Rakaa et al., 2025).

The findings of this study have important implications for inclusive education policies. Adapted physical activity programs should be part of the school curriculum, not only to improve PF but also to foster social integration and community participation. A recent meta-analysis shows that teachers’ attitudes are decisive for implementing inclusive practices (Tarantino et al., 2025). Therefore, our results reinforce the importance of educational and health policies that consider physical activity as a central component of an equitable and accessible curriculum.

Although the number of included studies was limited and heterogeneity was high, this work provides relevant evidence for teaching practice and educational policy. It highlights the need to implement adapted physical education programs and inclusive pedagogical strategies that promote active and equitable participation of students with hearing loss. Future research should therefore incorporate larger samples and longitudinal designs to provide greater precision and external validity.

## 5. Conclusions

This meta-analysis confirms that children and adolescents with HL perform significantly worse than their hearing peers in PF variables such as strength, agility, speed, and balance. These inequalities extend beyond biomedical aspects and directly affect school participation and educational equity.

Inclusive physical education and adapted activity programs are essential to close this gap and ensure equal opportunities. Incorporating inclusion and accessibility into the curriculum, together with teacher training in inclusive practices, can improve physical performance, self-esteem, and social integration. Therefore, the results of this study reinforce the importance of educational policies that recognize PE as a key component of an equitable and accessible curriculum.

Although the study has limitations, including small sample sizes and high heterogeneity, it makes an original contribution by integrating an inclusive perspective into the analysis of PF in children with hearing loss. This work provides a reference framework for future research and for designing adapted physical education programs that foster active and equitable participation.

## 6. Declarations

## 6.1. Acknowledgement

The authors would like to thank all researchers whose studies were included in this systematic review and meta-analysis.

## 6.2. Declaration of generative AI and AI-assisted technologies in the manuscript preparation process

During the preparation of this work, the authors used ChatGPT (Ope-nAI) to assist with language editing, text refinement, formatting, and consistency checks. After using this tool, the authors carefully reviewed, edited, and validated all content as necessary and take full responsibility for the content of the published article.

## 6.4. Ethics statement

Ethical approval and informed consent were not required for this study because it was based exclusively on data extracted from previously published studies.

## 6.5. Author contributions (CRediT)

María Victoria Díaz-Franco: Conceptualization, Methodology, Investigation, Data curation, Formal analysis, Writing – original draft, Writing – review & editing.

Alexis Caniuqueo-Vargas: Methodology, Investigation, Data curation, Formal analysis, Writing – review & editing.

Olusiji Adebola Lasekan: Formal analysis, Validation, Writing – review & editing.

Carlos Alberto Castillo Sarmiento: Conceptualization, Methodology, Supervision, Validation, Writing – review & editing.

Beatriz Rodríguez Martín: Conceptualization, Methodology, Supervision, Validation, Project administration, Writing – review & editing.

All authors read and approved the final manuscript.

## 6.7. Data Availability Statement

All supplementary data supporting the findings of this study have been included within the files uploaded to the journal’s submission platform. No additional datasets were generated or analyzed beyond those presented in the manuscript and supplementary materials.

## Appendix A. Supplementary material

Supplementary material related to this article is available online.

- Supplementary Material 1 provides the complete database search strategies.
- Supplementary Material 2 includes the standardized data extraction form used for study selection and data collection.

