## Supplementary material for "Inequalities in physical fitness in children with hearing loss: a systematic review and meta-analysis with implications for inclusive physical education and school curriculum": SM1

*Supplementary Material 1: Search command table for each database*

| ***Database*** | ***Search command*** |
|---|---|
| *PubMed* | *(((schoolchildren) OR (boys) OR (girls) OR (adolescents)) AND ((“physical condition”) OR (“Aerobic capacity”) OR (strength) OR (endurance) OR (speed) OR (Flexibility) OR (“physical ability”)) AND ((hearing) OR (“hearing loss”) OR (“hearing impairment”)))* |
| *Scopus* | *(((schoolchildren) OR (boys) OR (girls) OR (adolescents)) AND ((physical condition) OR (Aerobic capacity) OR (strength) OR (endurance) OR (speed) OR (Flexibility) OR (physical ability)) AND ((hearing) OR (hearing loss) OR (hearing impairment)))* |
| *Web of science* | *(((schoolchildren) OR (boys) OR (girls) OR (adolescents)) AND ((physical condition) OR (Aerobic capacity) OR (strength) OR (endurance) OR (speed) OR (Flexibility) OR (physical ability)) AND ((hearing) OR (hearing loss) OR (hearing impairment)))* |
