## Supplementary material for "Inequalities in physical fitness in children with hearing loss: a systematic review and meta-analysis with implications for inclusive physical education and school curriculum": SM2

*Supplementary Material 2: Studies retrieved from the literature search strategy.*

| Study | Participans<br>(n, sex, age, dg) | Physical Condition Performance Variable/ Test | Design | Test | Results/ Conclusions |
|---|---|---|---|---|---|
| Azadian 2020 | 30 children (15 with HL and 15 without HL; age 11,3 ± 1,8 and 10,5 ± 1,5 years) | Force based on the torque of the hip, knee, and ankle | Observational/ cross-sectional | Three-dimensional joint torques were analyzed during barefoot walking at preferred speed using Kistler force plates and a Vicon motion capture system. | Findings revealed that boys with hearing loss showed lower joint torques in ankle evertors, knee flexors, abductors and internal rotators as well as in hip internal rotators in both, the dominant and non-dominant lower limbs (all $p<0.05$). Further, in the dominant limb, larger peak ankle dorsiflexor ($p<0.001$), knee adductor ($p<0.001$), and hip adductor torques ($p<0.001$). Were found in deaf participants compared with controls. |
| Banerjee 2022 | 252 children (126 men and 126 woman) 42 without HL and 210 with HL; age 13 – 20 years. | Agility was measured through 4×10 meter shuttle run test | Observational/ cross-sectional | 4×10 meter shuttle run test | The significance was tested at $p <.05$ level. In agility, boys were significantly better than the girls in all respect, when different hearing impaired groups were combined. Significant differences in agility among the different hearing impaired groups were observed except in few cases when both sexes were combined. An increasing linear trend in agility was observed with decreasing degree of hearing Loss. Thus, an inverse relationship between agility and degree of hearing loss (dB HL) is observed. |
| Majlesi 2014 | 20 children (10 HL and 10 without HL; age 11,3 ± 1,9 and 10,4 ± 4,4 | Walking speed and balance (swing speed) | Experimental | Walking with the test Gait measurement. A four camera Vicon system (Oxford Metrics, Oxford, UK and Static balance with the test of Balance measurement. The static balance function of the subjects was measured by Kistler Force Platform (Type 9281, Kistler Instrument AG, Winterthur, Switzerland) at a frequency of 200 Hz.) | A comparison between the control and experimental groups revealed that the intervention program had not significantly increased gait velocity while it had significantly decreased the amount of sway. Thus, it was concluded that an exercise program that enhances somatosensory ability can result in improved balance in deaf children. |

| | | | | | |
|---|---|---|---|---|---|
| Melo 2012 | 88 children (44 with HL and 44 without HL; age 7 - 17 years) | Walking and balance | Cross-sectional | Balance and gait characteristics were performed using the Tinetti Balance and Mobility Scale and the gait velocity with the test Timed Up and Go | The results from the balance evaluation did not show significant differences between groups, genders or age groups; however, the deaf scholars had worse performance on clinic balance in all categories. In the evaluation of gait characteristics, there were significant differences between groups ($p<0.001$), genders ($p<0.001$), and age groups: 7–10 years-old ($p=0.022$) and 11–17 years-old ($p<0.001$). With respect to gait speed, results showed significant differences between groups only for female students ($p=0.027$) aged 7–10 years-old ($p<0.001$). |
| Shavel 2021 | 94 children (72 with HL and 22 without HL; age 6 – 10 years) | The physical condition of the schoolchildren was determined using a set of medical and biological methods: blood pressure measurement, heart rate calculation according to the electrocardiogram R-R interval, spirometry, electrocardiography, echocardiography, physical work capacity (PWC150) test, measurement of catecholamines using F. | Experimental | Physical work capacity (PWC150) test | A decrease in physical performance was found in deaf children compared to their normal-hearing peers for both age groups: girls – $295.97 \pm 4.26$ kg-m/min) ($p < 0.05$) and $310.37 \pm 2.69$ kg-m/min, respectively ($p < 0.05$); boys – $306.92 \pm 1.28$ kg-m/min and $315.07 \pm 1.39$ kg-m/min, respectively ($p < 0.05$). The results of the study on physical performance of deaf children aged 8-10 years old were significantly different from the results of children without hearing loss ($p <0.05$). |
